# Optical Genome Mapping Identifies a Novel Pediatric Embryonal Tumor Subtype with a *ZNF532-NUTM1* Fusion

**DOI:** 10.1101/2022.05.20.22275344

**Authors:** Miriam Bornhorst, Augustine Eze, Surajit Bhattacharya, Ethan Putnam, M. Isabel Almira-Suarez, Christopher Rossi, Madhuri Kambhampati, Miguel Almalvez, Joyce Turner, John Myseros, Eric Vilain, Roger J. Packer, Javad Nazarian, Brian Rood, Hayk Barseghyan

## Abstract

Molecular characteristics of pediatric brain tumors have not only allowed for tumor subgrouping but have introduced novel treatment options for patients with specific tumor alterations. Therefore, an accurate histologic and molecular diagnosis is critical for optimized management of all pediatric patients with brain tumors, including central nervous system embryonal tumors. We present a case where optical genome mapping identified a *ZNF532-NUTM1* fusion in a patient with a unique tumor best characterized histologically as a central nervous system embryonal tumor with rhabdoid features. Additional analyses including immunohistochemistry for NUT protein, methylation array, whole genome, and RNA-sequencing was done to confirm the presence of the fusion in the tumor. This is the first description of a pediatric patient with a *ZNF532-NUTM1* fusion, yet the histology of this tumor is similar to that of adult cancers with *ZNF-NUTM1* fusions and other *NUTM1-*fusion positive brain tumors reported in literature. Although rare, the distinct pathology and underlying molecular characteristics of these tumors separate them from other embryonal tumors. Therefore, the *NUTM-*rearrangement appears to define a novel subgroup of pediatric central nervous system embryonal tumors with rhabdoid/epithelioid features that may have a unique response to treatment. Screening for a *NUTM1-*rearrangement should be considered for all patients with unclassified central nervous system tumors with rhabdoid features to ensure accurate diagnosis so this can ultimately inform therapeutic management for these patients.

## Introduction

Embryonal tumors are a class of highly aggressive and heterogenous malignant central nervous system (CNS) tumors that primarily occur in infants and young children [1]. They are characterized histologically as undifferentiated or poorly differentiated tumors of neuroepithelial origin and have high cellular activity and rapid growth. There are several types of embryonal tumors including medulloblastoma and other embryonal tumors including atypical teratoid rhabdoid tumor (AT/RT) and embryonal tumors with multilayered rosettes (ETMR)[2,3]. To assist in distinguishing between the different classifications of embryonal tumors, genetic characterization, which has rapidly become part of the diagnosis workflow, is essential. Medulloblastoma is further subdivided into four categories: WNT-activated, SHH-activated, Group 3 (non-WNT/non-SHH), and Group 4 (non-WNT/non-SHH)[4]. ETMR tumors, which are rare and generally arise in infants, are mostly classified through the presence of multilayered neuroepithelial cells that resemble rosettes with or without the presence of C19MC alteration[5]. For AT/RTs, SMARCB1 (also called INI1) or less commonly SMARCA4 (BRG1) loss of function is a criterion for diagnosis[6]. CNS neuroblastoma, FOXR2-activated and CNS tumor with BCOR internal tandem duplication are embryonal tumors that have recently been added to the 2021 WHO classification guidelines based on their molecular signatures[3]. Tumors that have histologic features of AT/RT, but do not have either INI1 or BRG1 loss were classified as CNS embryonal tumors with rhabdoid features based on the 2016 WHO guidelines, and now are included in the CNS embryonal tumors, NOS category [1–3]. The tumors in this last subgroup do not have known/common molecular features.

Standard of care therapy for patients with embryonal tumors generally includes a combination of surgery, chemotherapy and radiation therapy[7–9]. Historically, therapeutic management primarily depended on patient characteristics such as age, tumor location, presence of metastatic disease and tumor histology (i.e. medulloblastoma vs AT/RT, etc). More recently, molecular characteristics of the tumors have introduced novel treatment options for patients with specific tumor alterations. For example, hedgehog pathway inhibitors are being included in clinical trials for patients with SHH-activated medulloblastoma, lower radiation doses are being trialed in patients with lower-risk WNT-activated medulloblastoma, and intensified therapies are being trialed in patients with high-risk embryonal tumors[9]. An accurate histological and molecular diagnosis of embryonal tumors is therefore very important for both risk stratification and treatment planning.

In this case, the patient presented with an unusual tumor that had histopathologic features best aligning to a CNS embryonal tumor with rhabdoid features. Optical genome mapping (OGM) revealed a novel *ZNF532-NUTM1* fusion that has not been described previously in children with any cancer type or adult brain tumor but has been identified in adults with tumors of the lung, mandible, parotid gland, and pelvic bone that have similar histologic features as the patient’s tumor. This case report describes clinical and genomic features of a patient with this novel pediatric embryonal tumor subtype.

### Case presentation

The patient was a male between 0-5 years of age and presented to care with stalled development, drooling, left-sided face flushing, left-sided eyelid drooping and daily complex partial seizures. MRI imaging revealed a cystic mass with an enhancing nodular component measuring 11×13 cm in the temporoparietal region (Figure 1A&B). Diffusion weighted imaging was consistent with a cellular lesion. MRI of the spine was negative for metastatic disease. A gross total resection (GTR) was performed without complication three days later (Figure 1C). On pathology, the tumor was a high-grade neoplasm with focal necrosis (Figure 1D). Undifferentiated cells with an embryonal morphology were the primary cell type, but there were also slightly larger cells with epithelioid or rhabdoid morphology (Figure 1 E&F). Mitotic figures were noted to be abundant (Figure 1E) and the tumor had a high proliferative index (Figure 1G). Immunohistochemistry (IHC) showed patchy GFAP, EMA and cytokeratin staining. p53 IHC staining was consistent with wild type and INI1 was retained (Figure 1H). Further IHC analysis showed retained BRG1, which ruled out an AT/RT. The patient was diagnosed with a WHO Grade IV CNS embryonal tumor with rhabdoid features based on the 2016 classification criteria[2].

**Figure 1:**
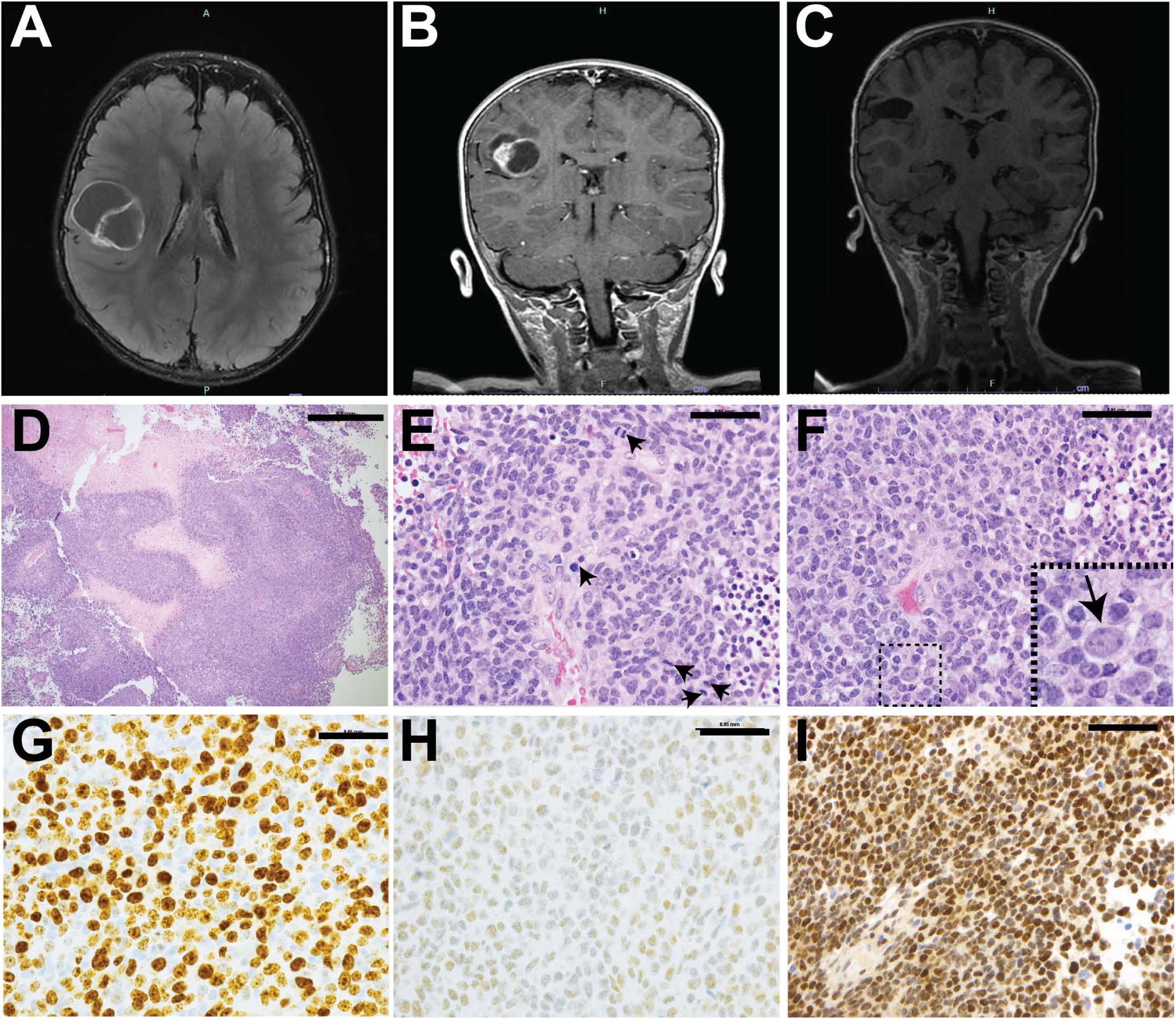
MRI and IHC images. Axial T2+contrast **(A)** and coronal T1+ contrast (**B)** MRI imaging showed an enhancing supra-insular/inferior parietal mixed cystic/nodular neoplasm. The lesion was completely removed with surgical resection **(C). (D)** H&E staining revealed a high-grade tumor organized in sheets (purple) with areas of necrosis (pale pink) (scale bar = 0.5mm). (E) Mitotic figures were abundant (arrow heads; scale bar = 0.05mm). (F) The undifferentiated tumor cells had an embryonal morphology and scattered larger cells with a vague epithelioid or rhabdoid morphology (arrow in inset shows an epithelioid-like cell; rhabdoid not seen in picture; scale bar = 0.05mm). Ki67 proliferative index was more than 90% (**G**; scale bar 0.05mm). **(H)** INI1 was retained on IHC. **(I)** NUT antibody IHC (NeoGenomics) showed strong nuclear staining of NUT protein (scale bar = 0.1mm).

The patient was treated with standard infant embryonal tumor chemotherapy (Cisplatin, Vincristine, Etoposide, Vincristine, Cyclophosphamide, Methotrexate) followed by 3 cycles of high-dose consolidation chemotherapy with stem cell rescue (Carboplatin, Thiotepa and Etoposide). The patient had good response to the induction chemotherapy with no evidence of disease noted on the pre-consolidation MRI. During consolidation cycle #1, the patient developed hypotension and veno-occlusive disease of the liver. Despite maximum intervention, the patient passed away approximately one week later.

Because of the patient’s early presentation with a unique tumor, the cancer genetics team was consulted to rule out possible germline cancer predisposition. Family history was significant for a sibling who died in utero, and distant history of leukemia, cervical cancer, colon and breast cancer on the maternal side of the family. A germline tumor panel including approximately 120 genes was performed and revealed a variant of unclear significance in the *MC1R* gene and a maternally inherited *PTCH1* gene mutation that was initially a variant of unclear significance but has subsequently been reclassified as a likely benign variant. Neither of these germline gene mutations were thought to be associated with his tumor development.

#### Molecular characterization

Tumor analysis was performed on a research basis and included Epic methylation array, Optical Genome Mapping, whole genome sequencing, and RNA sequencing (See supplementary file for methods). The EPIC methylation array results were entered into the tumor methylation classifier (MolecularNeuropathology.org) but the tumor did not cluster with any of the commonly diagnosed embryonal tumors (not classifiable) [10]. Whole genome sequencing analysis for single nucleotides and small insertions/deletions did not reveal significant Tier1 or Tier 2 somatic mutations. The tumor mutation burden was calculated to be <1 mut/Mb (low) and a majority of the variants identified were noted to be intronic.

Optical genome mapping (OGM; Bionano Saphyr Instrument), which utilizes ultra-long DNA molecules to assess for structural variants, showed complex three-way rearrangements amongst chromosomes 12, 18 and 15 in the tumor (Supplementary Figure 1). The most clinically significant event was an insertion that resulted in the fusion of *ZNF532* on chromosome 18 with *NUTM1* on chromosome 15 (Figure 2A). Additional rearrangements without clear clinical significance included (1) a translocation between chromosome 15 and 18; (2) an inversion on chromosome 18 near the same breakpoint as the translocation; (3) copy number variant change on chromosome 15 and (4) a small insertion on chromosome 18 derived from chromosome 12 (Supplementary Figure 2). Subsequent SV analysis of the whole genome sequencing data and additional RNA-sequencing confirmed the presence of the *ZNF532-NUTM1* fusion in the tumor (Figure 2B&C). No other clinically significant SVs or fusions were identified.

**Figure 2:**
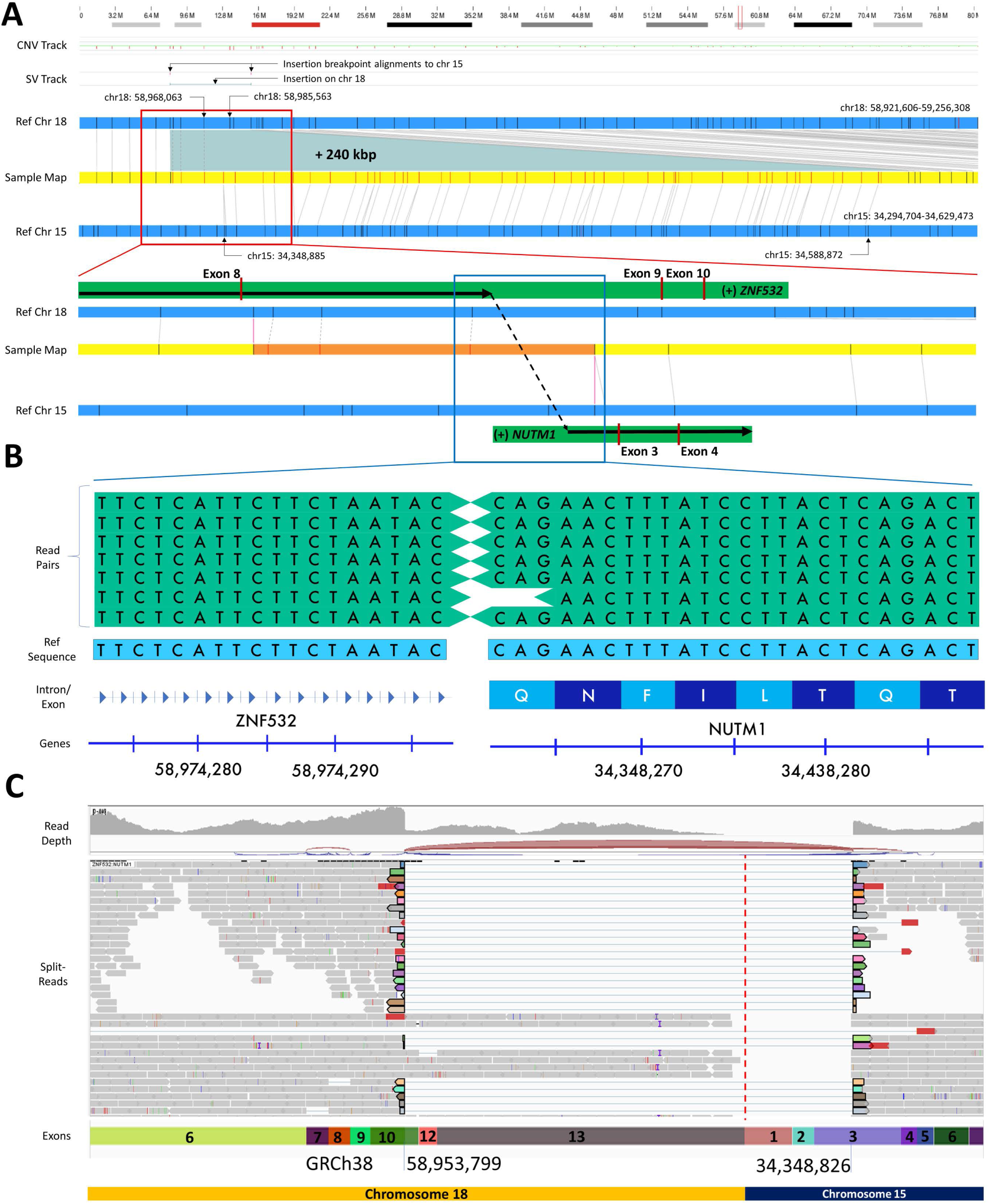
*ZNF532-NUTM1* fusion identified by optical genome mapping and confirmed by short-read sequencing. **(A)** OGM genome browser view of the identified fusion. Top: G-band staining of chromosome 18, followed by copy number and structural variant tracks. The green line and adjacent purple dots indicate the location of the insertion and corresponding breakpoints aligning to chromosome 15. The reference chromosomes 18 and 15 are shown in blue, with black vertical lines showing the DLE1 label locations. The assembled sample map is displayed in yellow, the red labels in the middle don’t have alignment to chromosome 18, instead they align to chromosome 15. The total insertion size from chromosome 15 to 18 is approximately 200kb. The left breakpoints on both chromosomes is magnified to show annotations of *ZNF532* and *NUTM1* genes, approximate breakpoint location and exons fused to exons. **(B)** Short-read sequencing alignments to the breakpoint location indicated by OGM. Read-pairs maps to two different chromosomes (chr18 and chr15), to confirm the identified translocation between an intron near *ZNF532* and exon 3 on *NUTM1* genes, respectively. **(C)** RNA-sequencing alignments also confirm the exon-exon fusion between *ZNF532* (exon 10) and *NUTM1* (exon3). The split reads are designated by color, with part of the reads mapping to exon 10 of chromosome 18 and other part mapping to exon 3, chromosome 15. The red dotted lines differentiate the 2 chromosomes.

In order to better understand the role and frequency of *ZNF532-NUTM1* fusions in pediatric brain tumors, a literature review using Pubmed was performed with a series of search strings including *ZNF532-NUTM1, NUTM1* AND brain tumor, and *ZNF532* AND brain tumor. The search was limited to English language articles concerning human studies exclusively and with publication dates going back 15 years from March 2022. Five additional cases with a *ZNF-NUTM1* fusion (four with *ZNF532*, one with *ZNF592)* and five reports of patients with *NUTM1-*rearranged brain tumors were identified (Figure 3A and Table1). *NUTM1* fusions in brain tumors are rare but have been identified in children and adults with supratentorial small-cell tumors similar to our patient (Table 1) [11–14]. The specific *ZNF532-NUTM1* fusion identified in our patient has not been reported in a child or a patient with a brain tumor prior to this case. However, this has been associated with adult lung, mandible, parotid gland, and pelvic bone cancers that have round cell and/or undifferentiated epithelioid morphology with or without a rhabdoid cell component, comparable to our patient’s tumor (Figure 3A, Figure 1D-F) [15–19].

**Figure 3:**
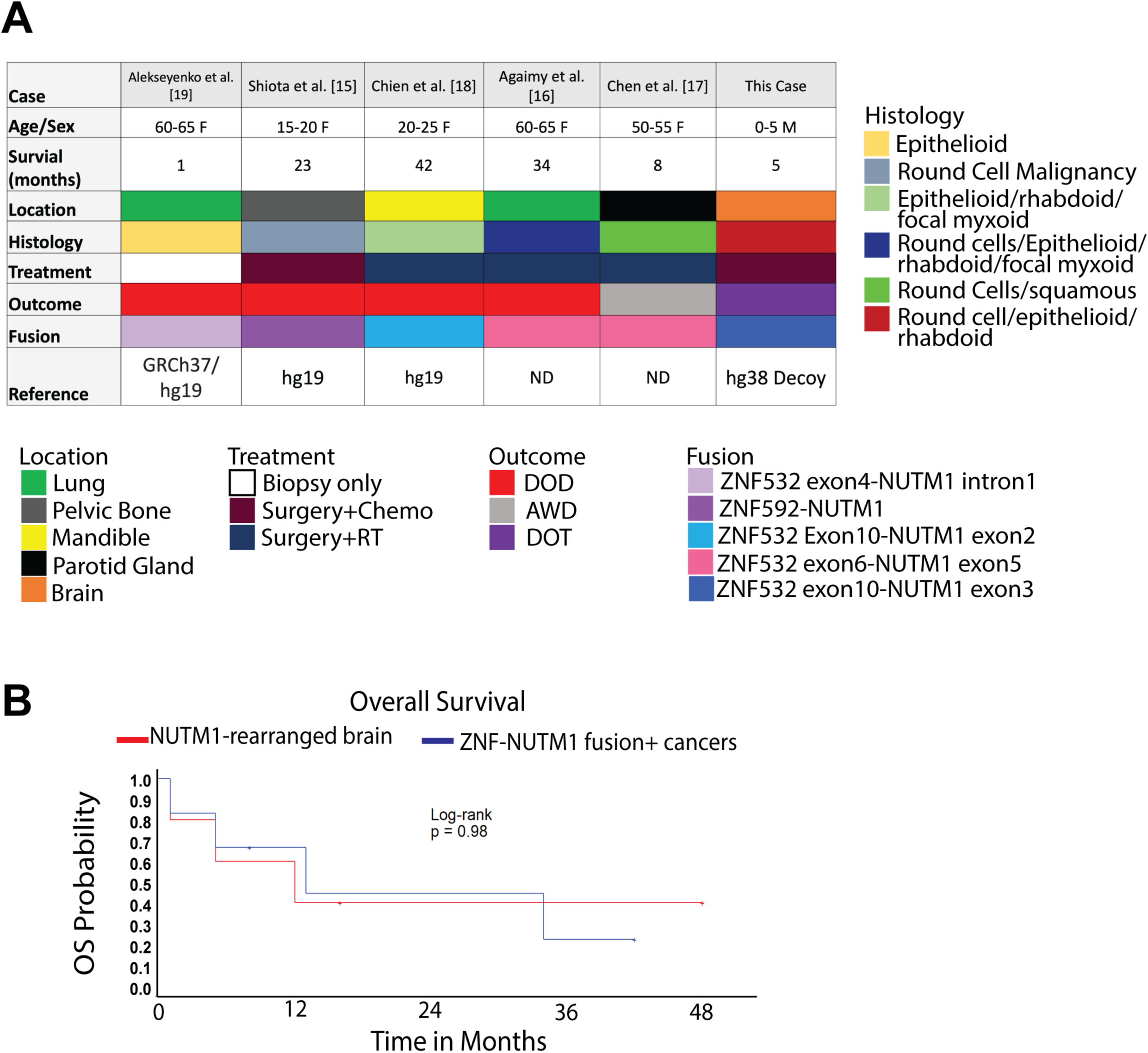
NUTM1 fusion tumors. **(A)** Oncoplot showing cancers with *ZNF-NUTM1* fusions. Ages are given as a 5 year range. **(B)** Kaplan-Meier curve showing overall survival of brain tumors with *NUTM1-* rearrangements (red line) and *ZNF-NUTM1* cancers (blue line).

**Table 1:** Previous cases from brain tumors with NUTM1 fusions.

| Reference | Age/<br>Sex | Site | Histology | IHC+ | Tx/<br>Outcome | Fusion |
| --- | --- | --- | --- | --- | --- | --- |
| Dickson<br>et al [11] | 0-5 yo<br>male | Left<br>parietal | Small round cells. Epithelioid-polygonal cells with a reticular-alveolar pattern and prominent myxoid stroma. Nuclear molding, speckled chromatin and conspicuous mitotic activity. | GFAP (2+, focal), synaptophysin (1+), NUT (5+). | Surgery, Chemo, DOD (12 mo) | *BRD4-NUTM1* |
| Sturm<br>et al [14] | 0-5 yo<br>female | Temporal/<br>Parietal | Small-cell phenotype, alveolar and fascicular growth | NUT (strong) | Unknown AWD (273 mo) | *CIC-NUTM1* |
| Sturm<br>et al [14] | 0-5 yo<br>female | Frontal/<br>Parietal | Small-cell phenotype, alveolar and fascicular growth | NUT (strong) | Unknown | *CIC-NUTM1* |
| Siegfried<br>et al [13] | 20-25<br>yo<br>female | Frontal | Fascicular architecture and chondro-myxoid areas; some neuron-like tumor cells; large nucleoli | NUT, GFAP (strong), p53, CD56. | Surgery, NED (16 mo) | *ATXN1-NUTM1* |
| Ko<br>et al [12] | 25-30<br>yo<br>female | Right<br>Frontal/<br>Temporal | Variegated tumor consisting mostly of small epithelioid cells with myxoid or fibrillar background | NUT, CD99, CD56, p53, GFAP (focal), neurofilament (focal). | Surgery, Chemo, DOD (1 mo) | *PARD3B-NUTM1* |
| This Case | 0-5 yo<br>male | Right<br>Temporal/<br>Parietal | Embryonal cell types, as well as epithelioid or rhabdoid-like cell types | GFAP (focal), Ki-67 high, NUT (strong), p53 wildtype | Surgery, Chemo, DOT (5 mo) | *ZNF532-NUTM1* |

GFAP indicates glial fibrillary acid protein; Chemo, chemotherapy; DOD, died of disease; AWD, alive with disease; NED, no evidence of disease; DOT, died of treatment. Ages are given as a 5 year range.

The patients reported in literature had a variety of different treatments. A multimodality treatment approach using surgery along with radiation, chemotherapy or both was most commonly employed, similar to the approach used with our patient. The one year overall survival was 40% and 45% in the *NUTM1-*rearranged brain tumor group and *ZNF532-NUTM1* cancer group respectively (Figure 3B).

An analysis of embryonal (n= 19) and AT/RT (n= 29) samples available through the Children’s Brain Tumor Network database did not reveal any additional tumors with a *ZNF532-NUTM1* fusion or a *NUTM1*- or *ZNF532-*rearrangement. However, this is a limited dataset and none of these tumors had pathology similar to the patient in this report. Additional analysis of a larger cohort of samples with round cell tumors and rhabdoid/epithelial features would be helpful to determine the frequency of these fusions in pediatric brain tumors.

## Discussion and Conclusions

Molecular characterization of pediatric brain tumors can help with both diagnostic and prognostic stratification. This has helped improve therapeutic strategies for patients with many tumors including embryonal tumors such as medulloblastoma and AT/RT. In this patient’s case, the tumor was described as an embryonal tumor with rhabdoid/epithelioid features that did not have genomic characteristics of an AT/RT and therefore was diagnosed under the umbrella term CNS embryonal tumor with rhabdoid features. Through optical genome mapping and subsequent whole genome and RNA sequencing, we uncovered a *ZNF532-NUTM1* mutation, which, based on previous reports in literature and the patient’s genomic and histologic findings, was most likely the genomic driver in this patient’s tumor.

*NUTM1*, is the *NUT* midline carcinoma gene family member 1 and located on chromosome 15q14[20]. *NUTM1-* rearranged tumors have a chromosomal translocation resulting in the fusion of the *NUTM1* gene on chromosome 15 with a gene involved in transcription regulation. The most common fusion partner is *BRD4* (chromosome 19) but fusions involving *BRD3* (chromosome 9), *NSD3* (chromosome 8) and zinc finger genes (*ZNF*) such as the *ZNF532* (chromosome 18) seen in our patient have also been described[20]. *NUTM1* rearrangements are most commonly associated with sarcomas and hematologic malignancies in both children and adults, although a variety of other cancers have also been identified. Children and infants with *NUTM1* fusions in B-cell ALL have favorable prognosis, while the clinical impact of this fusion on sarcomas is still unclear[20,21].

Although the oncogenic impact of the *ZNF532-NUTM1* fusion in CNS tumors has not been studied, functional studies of other cancers have shown that *NUTMI-*fusion positive cells demonstrate lack of differentiation of epithelial cells and nuclear localization of the NUT protein, suggesting that *NUTM1-*fusions contributed to tumor development by associating with nuclear chromatin and interfering with cell differentiation [22,23]. Consistent with this finding, nearly all of our patient’s tumor cells displayed homogenous nuclear expression of the monoclonal NUT antibody confirming nuclear localization of the NUT protein in this tumor (NeoGenomics; Figure 1I). *NUTM1-*rearranged tumors have also been described as having a lower mutation burden than other cancers, with an abundance of intronic mutations that do not affect canonical oncogenes or tumor suppressor genes[24,25]. This is consistent with the patient’s whole genome sequencing results that primarily showed intronic mutations with a low mutation burden and no other Tier1 or Tier2 pathogenic mutations.

Standard therapy for pediatric embryonal tumors includes a combination of surgery, radiation therapy and chemotherapy. The patient presented in this report had GTR followed by three cycles of induction chemotherapy and then one cycle of high dose chemotherapy followed by stem cell rescue before passing away from treatment complications. The patient had no radiographic evidence of disease at the time of passing. Upon review of CNS cases with *NUTM1-*fusions (Table 1) and patients with *ZNF532-NUTM1* associated cancers (Figure 3A), most patients had similar multi-modality treatment including surgery, RT and chemotherapy. Surgery, with the goal for GTR and negative margins, was performed when possible and has been shown to have a positive impact on overall outcomes in some patients with midline (head and neck) *NUT-*rearranged tumors [26–28]. Radiation therapy has also been shown to have some benefit in patients with *NUTM1-* rearranged midline tumors and has primarily been used in older patients with localized disease[26]. Chemotherapy has had mixed responses and thus far has not been demonstrated to improve overall outcomes in patients with these tumors[20,27]. However, a standardized therapeutic approach has not yet been identified in children or adults with *NUT-*rearranged tumors, so additional studies would be required to fully understand the role of surgery, radiation therapy, and chemotherapy in patients with these tumors.

In this report, we describe a novel pediatric brain tumor subtype with *NUTM1-*fusion positive pediatric CNS tumors, including the *ZNF532-NUTM1* fusion associated with rhabdoid/epithelial features on pathology. Although these tumors are rare, their distinct pathology and underlying molecular characteristics will likely separate them from other embryonal tumors in terms of response to treatment and targeted therapeutic options. Screening of CNS embryonal tumors that cannot be classified as medulloblastoma, AT/RT, ETMR or another molecular subgroup for the NUT protein through IHC followed by SV analysis either though WGS, RNA-sequencing or OGM to identify the specific *NUT-*rearrangement would be helpful for the identification of tumors that fit within this subgroup. Additional studies are needed to develop the best therapeutic options for these patients.

## Methods

### Sample processing

Ultra-high molecular weight (UHMW) DNA was extracted following manufacturer’s guidelines (Bionano Genomics Inc, USA) from flash frozen brain regions (superior frontal gyrus and primary visual cortex) as well as pelleted frozen PBMCs. Briefly, a total of 15-20mg of brain tissue or 1.5-2 million PBMCs were homogenized in cell buffer and digested with Proteinase K. DNA was precipitated with isopropanol and bound with nanobind magnetic disk. Bound UHMW DNA was resuspended in the elution buffer and quantified with Qubit dsDNA assay kits (ThermoFisher Scientific). Total RNA was extracted using Qiagen RNeasy Kit following manufacturer’s guidelines (Qiagen, Germany). RNA sequencing was performed at Novogene Inc, mRNA was selected from total RNA using poly-T oligo-attached magnetic beads and sequenced on an Illumina short-read instrument with 85 million reads (Novogene, USA).

DNA labeling was performed following manufacturer’s protocols (Bionano Genomics, USA). Direct Labeling Enzyme 1 (DLE-1) reactions were carried out using 750 ng of purified UHMW DNA. Labeled DNA was loaded on Saphyr chips for imaging. The fluorescently labeled DNA molecules were imaged sequentially across nanochannel arrays (Saphyr chip) on a Saphyr instrument (Bionano Genomics Inc, USA). Effective genome coverage of greater than 500X was achieved for all samples. All samples also met the following QC metrics: labelling density of ∼15/100 kbp; filtered (>15kbp) N50 > 230 kbp; map rate > 70%.

### Optical genome mapping analysis

Genome analysis was performed using software solutions provided by Bionano Genomics Inc. Automated, OGM specific, pipelines – Bionano Access and Solve (versions 1.7 and 3.7, respectively), were used for data processing and variant calling. **De novo assembly** was performed using Bionano’s custom assembler software program based on the Overlap-Layout-Consensus paradigm. Pairwise comparison of all DNA molecules was done to generate the initial consensus genome maps (*.cmap). Genome maps were further refined and extended with best matching molecules. SVs were identified based on the alignment profiles between the *de novo* assembled genome maps and the Human Genome Reference Consortium GRCh38 assembly. If the assembled map did not align contiguously to the reference, but instead were punctuated by internal alignment gaps (outlier) or end alignment gaps (endoutlier), then a putative SV was identified. **Rare variant analyses** were performed to performed to capture mosaic SVs occurring at low allelic fractions. Molecules of a given sample dataset were first aligned against GRCh38 assembly. SVs were identified based on discrepant alignment between sample molecules and reference genome, with no assumption about ploidy. Consensus genome maps (*.cmaps) were then assembled from clustered sets of molecules that identify the same variant. Finally, the cmaps were realigned to GRCh38, with SV data confirmed by consensus forming final SV calls. **Fractional copy number** analyses were performed from alignment of molecules and labels against GRCh38 (alignmolvrefsv). A sample’s raw label coverage was normalized against relative coverage from normal human controls, segmented, and baseline CN state estimated from calculating mode of coverage of all labels. If chromosome Y molecules were present, baseline coverage in sex chromosomes was halved. With a baseline estimated, CN states of segmented genomic intervals were assessed for significant increase/decrease from the baseline. Corresponding copy number gains and losses were exported. Certain SV and CN calls were masked, if occurring in GRC38 regions found to be in high variance (gaps, segmental duplications, etc.)

### Variant analysis

Bionano Access (Bionano Genomics Inc, USA) was used for SV annotation and filtering. Variants were filtered in access and nanotatoR [29] based on the following criteria: for *de novo*, rare variant and CNV pipelines, SVs were filtered based on Bionano Genomics recommended size and confidence cutoff values (e.g., >500bp/5kbp size cutoff for *de novo* assembly and rare variant pipelines respectively for INDELs). Rare SVs were selected by filtering out common variants with population frequency of >1% using Bionano Genomics’ database of SVs containing >300 healthy individuals. To select for potential clinically significant aberrations a gene list overlapping SVs was used.

### Genome sequence analysis

FASTQ reads were aligned to GRCh38 reference genome using, BWA-MEM [30], followed by processing of the aligned bam (for variant calling) using SAMtools [31] and Picard (Broad Institute). Next, for small nucleotide variant (SNV) and small insertion and deletion (INDEL) calling we use Mutect2 [32], followed by annotation using ANNOVAR [33]. Due to absence of a non-tumor tissues from the same individual, we used a 1000 genome panel of normal (PON) variant call file from Broad Institute, as a proxy. Quality filtration for SNV/INDEL was performed using the Mutect2 function. *FilterMutectCalls*. For larger structural variant calls, we used Manta [34], followed by annotation using AnnotSV [35]. For the SV visualization Integrative Genome Viewer (IGV) was used.

### RNA-sequence analysis

RNA-seq data was aligned to GRCh38 reference genome, followed by fusion calling using STAR-Fusion [36]. Visualization of the fusion was performed using Clinker [37] and IGV.

### EPIC methylation chip analysis

An input of 300 ng of DNA was bisulfite-converted using the DNA Methylation-Lightning kit (Zymo Research). After whole-genome amplification and enzymatic fragmentation, samples were hybridized to BeadChip arrays using the Infinium Methylation EPIC BeadChip kit according to the manufacturer’s protocol (Illumina). Intensity values at the over 850,000 methylation sites on the BeadChips were measured across the genome at single-nucleotide resolution using iScan (Illumina). For classifying the tumors, CNS tumor classification tool hosted at molecularneuropathology.org, was used on the methylation signal files [10].

## Data Availability

All data produced in the present study are available upon reasonable request to the authors

## Declarations

### Ethics approval and consent to participate

Written informed consent was obtained from the parents of the patient through IRB approved Children’s National Hospital protocol, Pro#1339, PI Javad Nazarian, PhD.

### Consent for publication

The participant’s family provided consent for submission of the case for publication.

### Availability of data and materials

Results of all data generated or analysed in this study are either available in the published manuscript or available upon reasonable request. The datasets (including raw data files) used for data analysis during the current study are available from the corresponding author on reasonable request.

### Authors’ contributions

HB, MB and SB analyzed and interpreted the patient genomic data. MB and HB prepared figures and wrote the manuscript. AE was a major contributor to writing the manuscript, the literature search and figure preparation. EP performed literature search and assisted with the figures. MIAS and CR assisted with the pathology interpretation and figures. MK performed the EPIC array methylation analysis. MA assisted with DNA and RNA extraction and analysis. JT, JM, RP and BR participated in patient care. EV and JN provided mentorship with data analysis. All authors read and approved the final manuscript.

## Acknowledgements

We would like to acknowledge the patient’s family who donated tissue for research.

## Figure legends

**Supplementary Figure 1:**
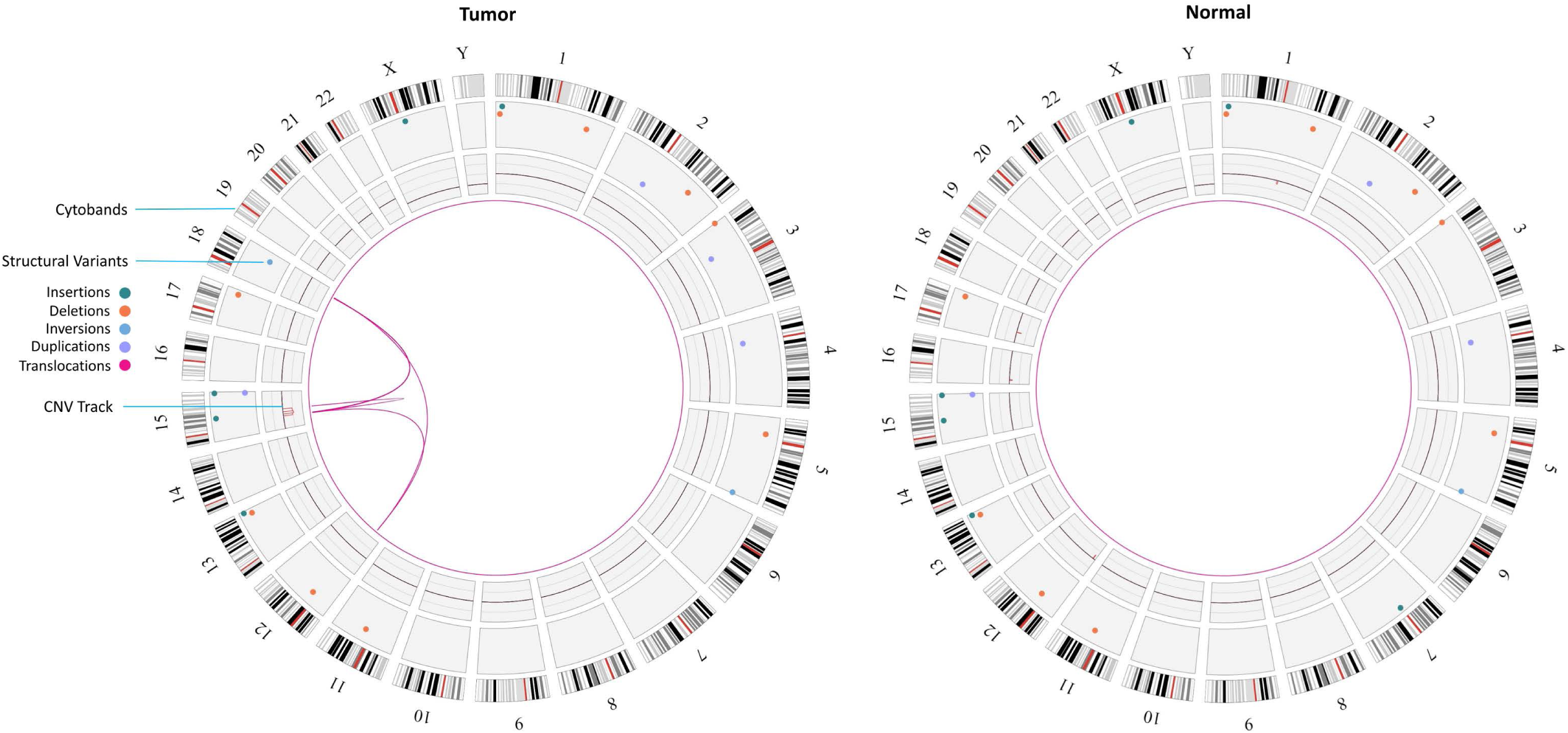
Representation of the SVs identified in the case in tumor tissue sample and control blood sample. Four level circos plots displaying chromosomal G-band on the outer level, SVs in the middle (insertions – green, deletions – orange, inversions – cyan, duplications – purple, translocation – purple), followed by CNV track (gains – blue, loss – orange) and inner most circle displaying the translocation connections between the chromosomes.

**Supplementary Figure 2:**
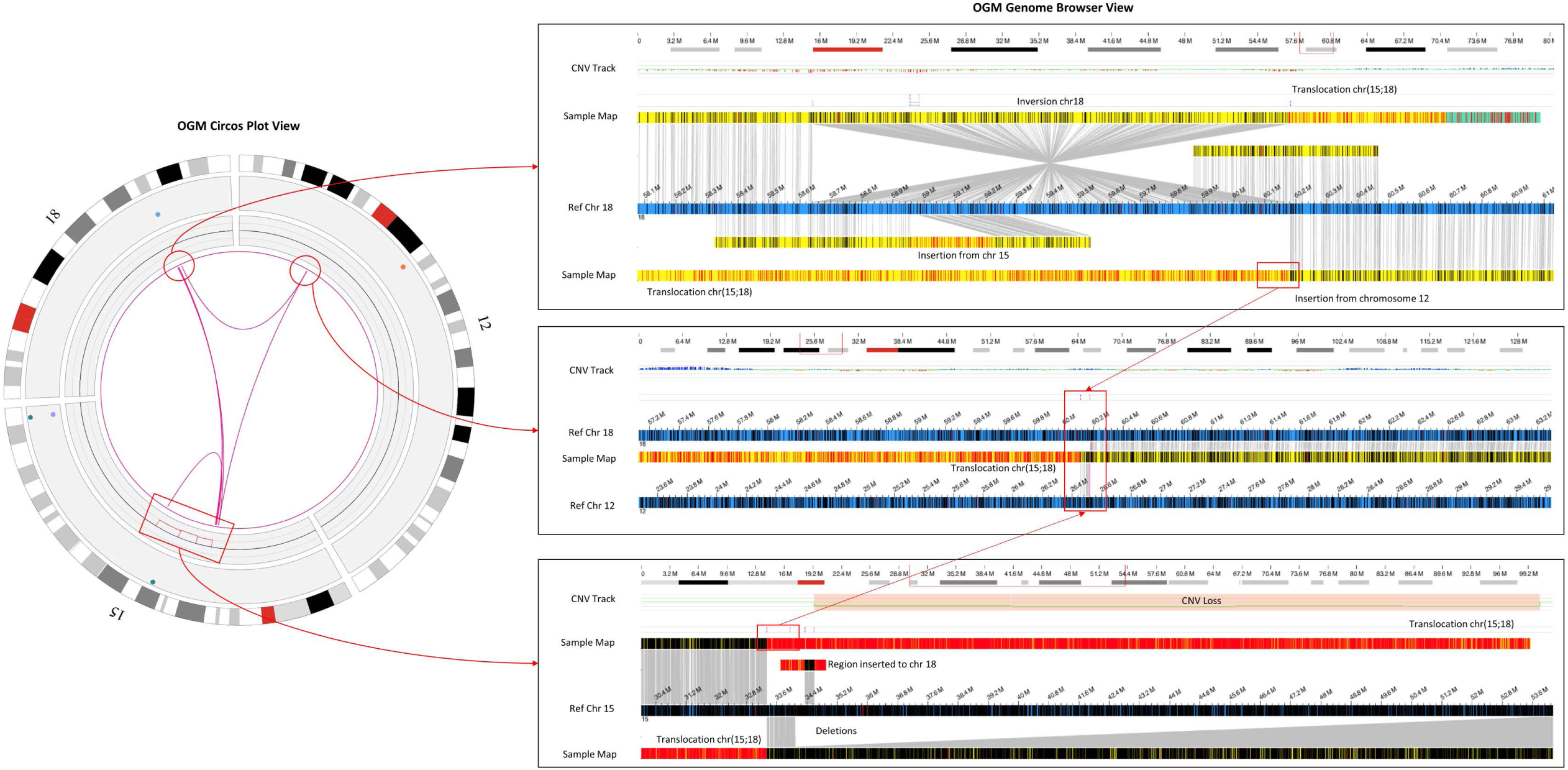
Complex rearrangements observed in the tumor sample. Left: Circos plot summarizing the rearrangements seen between chromosomes 12, 15 and 18 (purple connecting lines). Overall, the reference chromosomes are displayed in blue and the assembled sample maps in yellow. DLE1 label sites are in black. Top: Chromosome 18 genome view around the breakpoints between chromosome 12 and 15. An insertion from chromosome 15 and 12 is observed. Additionally, the chromosome 15 and 18 translocation junction results in an inversion as observed on chromosome 18. Middle: Alignments of labels from chromosome 12 to assembled maps on chromosomes 15 and 18, indicating that this DNA material has been inserted onto chromosome 18. Bottom: Translocations between chromosome 15 and 18 observed by two maps as well as a large CN loss on chromosome 15.

